# GUÍA: a digital platform to facilitate result disclosure in genetic counseling

**DOI:** 10.1101/2020.09.14.20194191

**Authors:** Sabrina A. Suckiel, Jaqueline A. Odgis, Katie M. Gallagher, Jessica E. Rodriguez, Dana Watnick, Gabrielle Bertier, Monisha Sebastin, Nicole Yelton, Estefany Maria, Jessenia Lopez, Michelle Ramos, Nicole Kelly, Nehama Teitelman, Faygel Beren, Tom Kaszemacher, Kojo Davis, Irma Laguerre, Lynne D. Richardson, George A. Diaz, Nathaniel M. Pearson, Stephen B. Ellis, Christian Stolte, Mimsie Robinson, Patricia Kovatch, Carol R. Horowitz, Bruce D. Gelb, John M. Greally, Laurie J. Bauman, Randi E. Zinberg, Noura Abul-Husn, Melissa P. Wasserstein, Eimear E. Kenny

## Abstract

**Purpose:** Use of genomic sequencing is increasing at a pace that requires technological solutions to effectively meet the needs of a growing patient population. We developed GUÍA, a web-based application, to enhance the delivery of genomic results and related clinical information to patients and families.

**Methods:** GUÍA development occurred in 5 phases: formative research, content development, user interface design, stakeholder/community member input, and web application development. Development was informed by qualitative research involving parents (N=22) whose children underwent genomic testing. Participants enrolled in the NYCKidSeq pilot study (N=18) completed structured feedback interviews post-result disclosure using GUÍA. Genetic specialists, researchers, patients, and community stakeholders provided their perspectives on GUÍA’s design to ensure technical, cultural, and literacy appropriateness.

**Results:** NYCKidSeq participants responded positively to the use of GUÍA to deliver their children’s results. All participants (N=10) with previous experience with genetic testing felt GUÍA improved result disclosure, and 17 (94%) participants said the content was clear.

**Conclusions:** GUÍA communicates complex genomic information in an understandable and personalized manner. Initial piloting demonstrated GUÍA’s utility for families enrolled NYCKidSeq pilot study. Findings from the NYCKidSeq clinical trial will provide insight into GUÍA’s effectiveness in communicating results among diverse, multilingual populations.

## INTRODUCTION

Rapid technological advancements in genomic sequencing (GS) in the past two decades have escalated the use of genomic data for diagnosing, predicting, and preventing disease, resulting in an increasing integration of genomic information in clinical decision-making. The shifting landscape of genetic testing has had a substantial impact on the practice of genetic counseling. Genetic counselors (GCs) are trained to help patients understand complex genomic information. Results from GS often reside along a spectrum of pathogenicity and can be either uninformative or ambiguous, while interpretation of results can change over time as evidence for pathogenicity accumulates. There is also the possibility of identifying findings unrelated to the primary purpose of the testing, and findings that may have peripheral impact upon family members, who are likely to have the limited genetics expertise of the general population.^1^ For all these reasons, genomic information is challenging to convey effectively. Despite this, GS is increasingly offered in a wide diversity of clinical settings, contributing to a greater demand for genomic medicine services across a variety of healthcare specialties.^2^ With the expansion of GS, GCs are challenged to scale services to meet the needs of a wide array of providers, patients, and their families.

Innovations in other technologies, including smartphones, artificial intelligence, and digital communication, are increasingly playing an important role in health systems.^3-5^ While genomic technology is driving the proliferation of inexpensive and accessible genetic tests, communication technology has also had a substantial impact on the practice of genetic counseling. Some technological solutions aim to bridge the barrier of access to counseling services. These include telehealth platforms, decision support tools, and genetic counseling aids such as educational videos and interactive web-based learning.^6^ Artificial-intelligence solutions like chatbots are emerging as tools to help patients navigate genetic testing.^7^ There are also self-service technology solutions designed for consumers and patients available through commercial laboratory websites that include self-guided educational modules, results delivery, and the option to speak with a GC.^8-10^ Other models like My46, a web-based tool that allows individuals to manage their genetic results, and Genomics ADvISER, an online, interactive decision aid to help patients in selecting secondary findings, have been born out of research initiatives to facilitate education and support genomic medicine implementation outside of clinic walls. ^7,11,12^

While these technological tools may help to improve access by streamlining the genetic counseling workflow, the majority of the patient-facing educational content is designed to address patients’ pre-test educational needs or tailored to consumer-driven genetic testing. These solutions are not designed to enhance the delivery of genomic results nor are they designed to increase understanding of the results and associated medical recommendations. Furthermore, there is a paucity of research on alternative delivery models and technological solutions for delivering genomic medicine and counseling services in diverse populations. Large scale, national research programs, such as the All of Us Research Program,^13^ the eMERGE Network,^14^ and the Clinical Sequencing Evidence-Generating Research (CSER) consortium, are exploring methods for implementing genomic medicine across diverse populations and settings.^15^ Outcomes from these studies will help to inform best practices for delivering genomic medicine services.

We created the Genomic Understanding, Information and Awareness (GUÍA, Spanish for ‘guide’) application, a novel web-based application designed to facilitate the delivery of GS results and related clinical information to participants and families of diverse backgrounds enrolled in the NYCKidSeq Project. GUÍA allows GCs to walk patients through their genomic test results in a personalized, highly visual, and narrative manner. GUÍA was developed based on the perspectives and input of providers, patients, and community stakeholders as part of the joint NHGRI and NIMHD-funded NYCKidSeq Project, a member of the CSER consortium. The development of GUÍA occurred in 5 discrete phases, including a formative qualitative study (phase 1), assembly of an expert working group and content development (phase 2), design of the user interface (phase 3), stakeholder and community member input (phase 4), and web application development (phase 5) (**Figure 1**). GUÍA was then piloted with 18 participants enrolled in the NYCKidSeq pilot phase.

**Figure 1.**
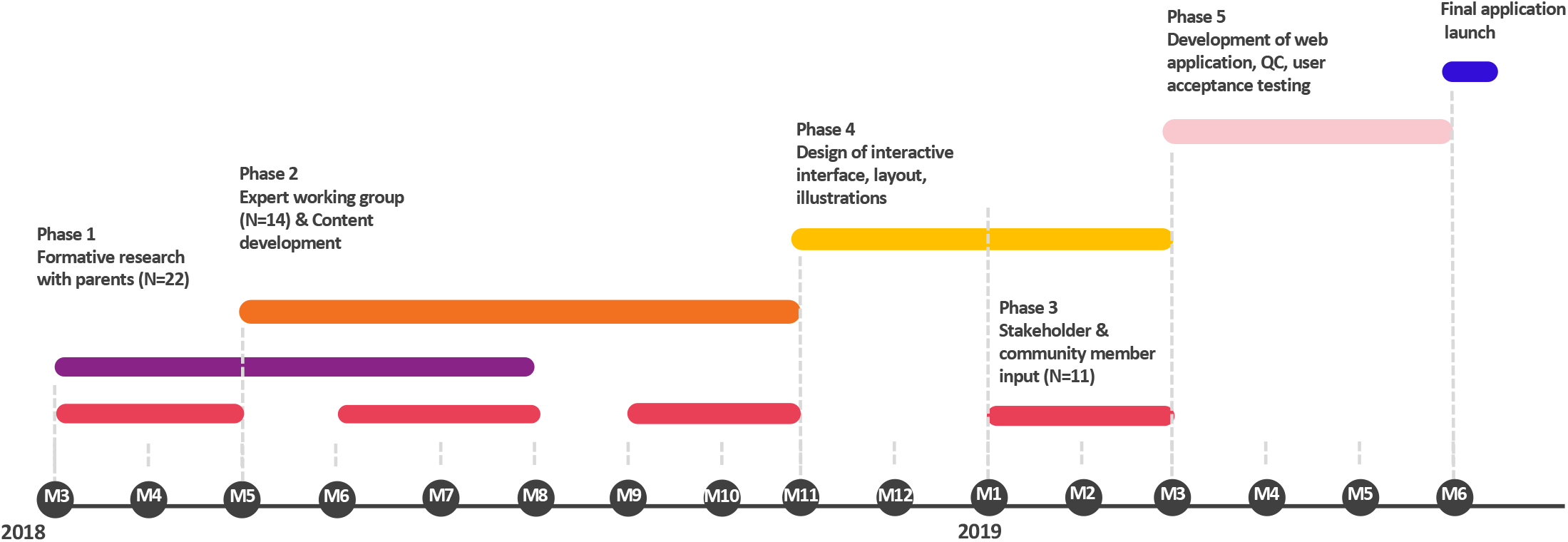
GUÍA development timeline.

## MATERIALS AND METHODS

### Formative Study

#### Setting and Study Population

We recruited parents, 20 mothers and 2 mother-father pairs, of 22 index children who had undergone exome sequencing, target gene panel, or microarray clinical genetic testing within the previous 12-months to participate in an in-depth interview. Participants were recruited at two major health systems in New York City: Mount Sinai Health System and Montefiore Medical Center. We used a stratified purposive sampling approach to recruit a diverse group of participants: 5 Black/African American (AA), 10 Hispanic/Latinx (H/L), 5 White/European American (EA), and 2 who self-reported as more than one race or ethnicity group (MR). We also stratified the sample to ensure representation of different testing results: 10 negative, 7 uncertain and 5 positive results.

#### Study Design

Parents received transportation costs and a $50 gift card for participating. In-depth interviews were conducted using narrative and focused interviewing techniques to highlight parents’ storied experiences paired with systematic probing for breadth and depth.^16,17^ A semi-structured interview guide was used to address the following domains: perceived purpose of testing; expectations of results; the return of results process and sequelae; and emotional responses. Interviews were administered in English or Spanish, were recorded, transcribed and translated into English.

#### Analysis

Analysis was conducted by a multidisciplinary team including GCs, a medical geneticist, and qualitative methods experts. First, individuals conducted case-based analyses to identify “repeating ideas” as a first step in codebook development.^17^ Then, team members independently applied the codebook to an interview to identify areas of (dis)agreement across coders, to develop code definitions and clarify labeling of emergent themes. Groups of two coders (1 GC + 1 non-GC) then applied the codebook to a single interview until consensus was achieved on current and new code applications. Using grounded theory’s constant comparative method^18,19^, this process was repeated until all interviews were coded by the 2-person groups. Full-team meetings were held to resolve issues and to discuss higher order theme development across the entire dataset.^19^ Findings were communicated to the Expert Working Group and the Genomics Community Board to inform GUÍA content development.

### Expert Working Group

An Expert Working Group (EWG) was formed with trans-disciplinary expertise in the development of education and medical genetics content for GUÍA. The group included four GCs, three medical geneticists, a pediatric cardiologist, a population geneticist, and a sociologist. The EWG led the development of the overall design of the application and the development of the content.

### Genomics Community Board

Members of the GUÍA development team met regularly with the Genomics Community Board (GCB), a subcommittee of the Mount Sinai community-academic research partnership board. The GCB is made up of community leaders, patients, and clinicians predominantly from Harlem and the Bronx who self-report as Black, African, African American, Afro-Latinx, or Afro-Caribbean.^20^ The GCB contributed to the conceptual design of GUÍA, reviewed development plans, prototypes and final versions of visuals and text throughout the development process.

### User Interface and Content Design of GUÍA

The EWG worked with experts in data visualization and user interface (UI) design to draft the design specifications for the application. The principal design specifications included: personalization (displaying name and preferred gender pronoun); text, illustrations, and hyperlinks on web pages; ability to display designated pages, data points, and illustrations based upon result type; tiered complexity of information; easy navigation; bilingual capability; option to export as pdf and print full content; compatibility for desktop and tablet. The UI designer produced the user experience framework which was designed for ease of navigation and ensured that all displayed content was relevant to the patient-participant’s result type. Using Zeplin^21^ collaborative design software, the EWG and the UI designer collaborated to produce a final set of wireframes (page schematics) and visual designs, which were used as the basis for the personalized result templates.

A bilingual version of each wireframe was generated that displayed both English (small font) and Spanish text (large font). We aimed to ensure that the Spanish text was understandable to patient-participants who spoke different dialects of Spanish, therefore, the translation team included individuals representing 5 Spanish dialects: Spanish, Mexican, Cuban, Dominican, and Puerto Rican. Discrepancies in the translated text were discussed and resolved.

### Development of the GUÍA Web Portal

The EWG worked with the software development team and UI expert to develop a specification for GUÍA development, and to establish the display logic and the data flow. The backend database contains a web form through which the GC inputs patient-participant specific information to populate the patient-facing frontend application. The database includes a library of template text including over 5,000 variables and approximately 50 illustrations that are coded to be displayed based on patient-participant specific results. A team of software engineers built the application over a 3-month period, followed by a 4-week User Assurance Testing period, during which the EWG performed extensive UI testing to evaluate UI interactions before the platform went live.

### Pilot Testing and Evaluation of GUÍA

#### Setting and Study Population

GUÍA was piloted in clinical settings with 18 participants enrolled in the lead-in phase of the NYCKidSeq clinical trial, which is described in detail elsewhere (Odgis et al. 2020, unpublished). Four GCs used GUÍA during result disclosures with participants/families. Details of each case were input as both structured and free text by the GCs prior to result disclosure. Structured text can be displayed in both English or Spanish, and free text can be inputted in both languages. Spanish free text is generated using Google Translate, and Spanish-speaking staff review the translations for accuracy. Feedback was solicited from the GCs regarding their experience using GUÍA.

#### Study Design and Analysis

We developed a brief, structured feedback guide to explore parents’ reactions to GUÍA. The guide contained 14 questions designed to capture general and specific reactions to GUÍA content and design elements. Research coordinators interviewed parents in English or Spanish directly following disclosure of their child’s genomic results using GUÍA. Feedback sessions were audio-recorded and parents were provided with a $40 gift card. A separate research coordinator reviewed the audio-recordings and extracted structured and narrative details of the participants responses. The responses were reviewed for themes relating to the GUÍA interface, including: layout, language, images, health literacy, and navigation.

### Ethics Statement

The Institutional Review Boards (IRB) of Icahn School of Medicine at Mount Sinai and Albert Einstein College of Medicine approved the Formative Study and the NYCKidSeq Study. Informed consent was obtained from all participants.

## RESULTS

### Phase 1 – Formative Study

The formative study comprised 22 interviews with parents of children who had received genomic results within the previous 12-months, sampling for diversity in both child race/ethnicity and type of genomic result. Seven core themes were identified, which are listed in **Table 1** with participant quotes exemplifying each theme.

**Table 1.**
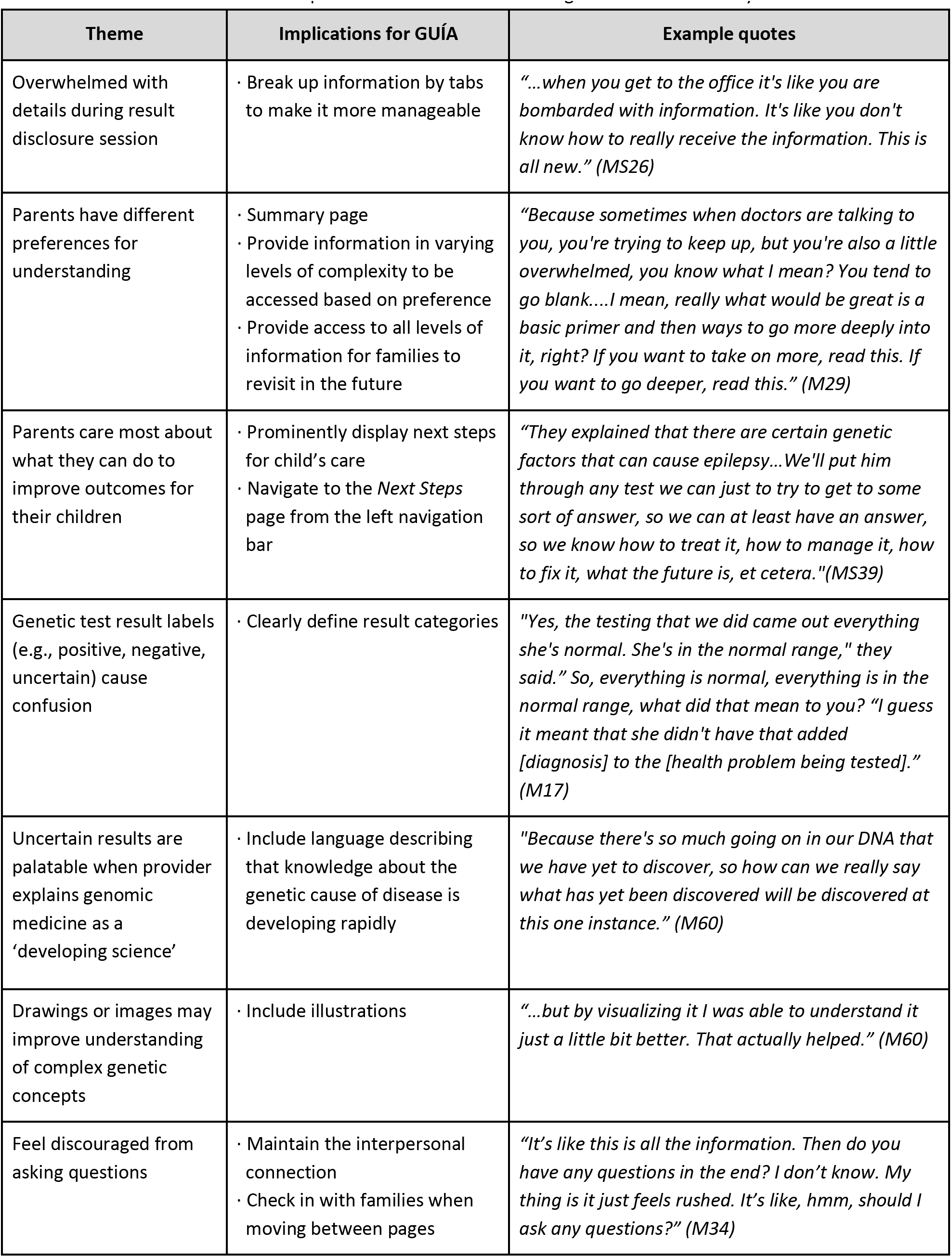
Themes related to the development of GUÍA identified through the Formative Study

One core theme is that parents often feel overwhelmed by the amount of information and details provided during a result disclosure session. There is considerable variability in what information parents want about genetics, genetic testing and the results, and in how much depth. This appears to be due to differences in the degree of understanding the parent feels comfortable with. To accommodate for a variety of educational preferences, we developed GUÍA to present information in increasing levels of complexity and on different web pages, so that patient-participants can modulate how many pages they access and the order in which they access these pages. For instance, for positive results, the overall, “big picture” details of the result are presented on the top page, and further genetic and condition details are displayed on separate tabs (**Supplemental Figure 1**; **Supplemental Table 1**). This allows the patient-participant to adjust the level of detail they receive in a GC session based on their personal preference.

Parents’ primary concern was improving health outcomes for their child. Consequently, parents were more interested in understanding how genetic results affected their child’s care than understanding technical details. We therefore designed GUÍA to prominently display follow-up care recommendations on the *Next Steps* page. Parents expressed confusion about the labels of genetic test results. For example, negative results were sometimes described as “normal” and were confused with the child’s symptoms. GUÍA clearly defines the result category (positive, uncertain, negative) on both the *Learn about Genome Sequencing* page and the *Result Summary* page.

Another theme identified was that parents who received uncertain results adapted to the uncertainty when the provider explained that genomic medicine is a developing science and knowledge of the genetic cause of disease will increase over time. We included language addressing this in GUÍA. Additionally, drawing complex concepts and showing illustrations to parents were appreciated. Parents expressed the desire to take home drawings made by a provider to revisit with family members. Thus, illustrations are integrated throughout the application, including on the education pages, the *Genetic Results* pages, and the *Inheritance* pages.

### Phase 2 – Content Development

The EWG first identified discrete components of a typical pediatric genetic counseling result disclosure session, as shown in the schema in **Supplemental Figure 1**. GUÍA addresses the following components: genetic education, primary (related to primary purpose of the test) and secondary (unrelated to the primary purpose of the test) results disclosure, clinical implications, inheritance and family implications, and resources. Contracting and psychosocial counseling are addressed through the interpersonal interaction between the counselor and the patient-participant.

To facilitate readability and comprehension, all text was written in the active voice and at the lowest possible reading level. We calculated the reading grade level by averaging the Flesch-Kincaid grade level of four randomly identified cases representing the three primary results categories (positive, negative, and uncertain) and a positive secondary finding result. While our goal reading level was 5^th^ grade, as recommended by the Joint Commission for patient education materials,^22^ the inclusion of genetic terminology meant that we could only decrease the reading level to an average 9^th^ grade level.

### Phase 3 – Stakeholder and Community Member Input

During the development of GUÍA, we met frequently with the Genomics Community Board (GCB) to discuss how a web-based application could be enhanced for the provision of genetic counseling for historically underserved populations, and to receive input on the content and design. Examples of specific implications for the application include: using illustrations inspired by biology textbooks to help explain complex concepts, ensuring images were culturally appropriate and resonated with the target audience, defining results categories clearly, simplifying pages by removing superfluous text, and including referrals to appropriate support services (see **Supplemental Table 2** GCB feedback).

### Phase 4-5 -Web Application Design and Development

We designed GUÍA to have a user-friendly interface with a visual design that is meant to make information communicated during the result disclosure easier to understand. GUÍA enables GCs to guide patients through the distinct components of a traditional result disclosure session while catering to patients’ diverse learning styles. Using GUÍA can be a counselor- or patient-driven experience and its modular design permits users to control the order in which information is accessed. If desired, the user can follow a proposed linear flow by using the navigation arrows at the bottom of each page, or they can navigate to different pages of the application from the *Home Page* or the navigation panel. The user can toggle between Spanish/English or English only text by clicking the language button. Illustrations accompanied by plain language text are incorporated throughout the application to convey key concepts, such as inheritance patterns, and demonstrate aspects specific to the patient-participant’s condition, such as affected organ systems.

The GUÍA sitemap (**Supplemental Figure 1**) displays the structure of the application, which consists of 9 distinct pages with sub-tabs to access educational modules, primary and secondary results, inheritance information and resource links. A high-level summary page provides key takeaways for the patient including results, next steps, and links to additional resources. **Figure 2** shows the GUÍA *Home Page* (a), and the *Result Summary* page (b) and *Family* page (c) presented in Spanish/English for a positive genetic result. The *Condition* and *Genetic Details* sub-tabs on the *Result Summary* page are personalized for the patient-participant and populated with relevant information garnered from the genetic test report and from the GC’s research into the patient-participant’s genetic condition. At the end of the counseling session, the entire GUÍA experience or individual pages can be converted to a PDF file format for the patient-participant’s use outside of the session.

**Figure 2.**
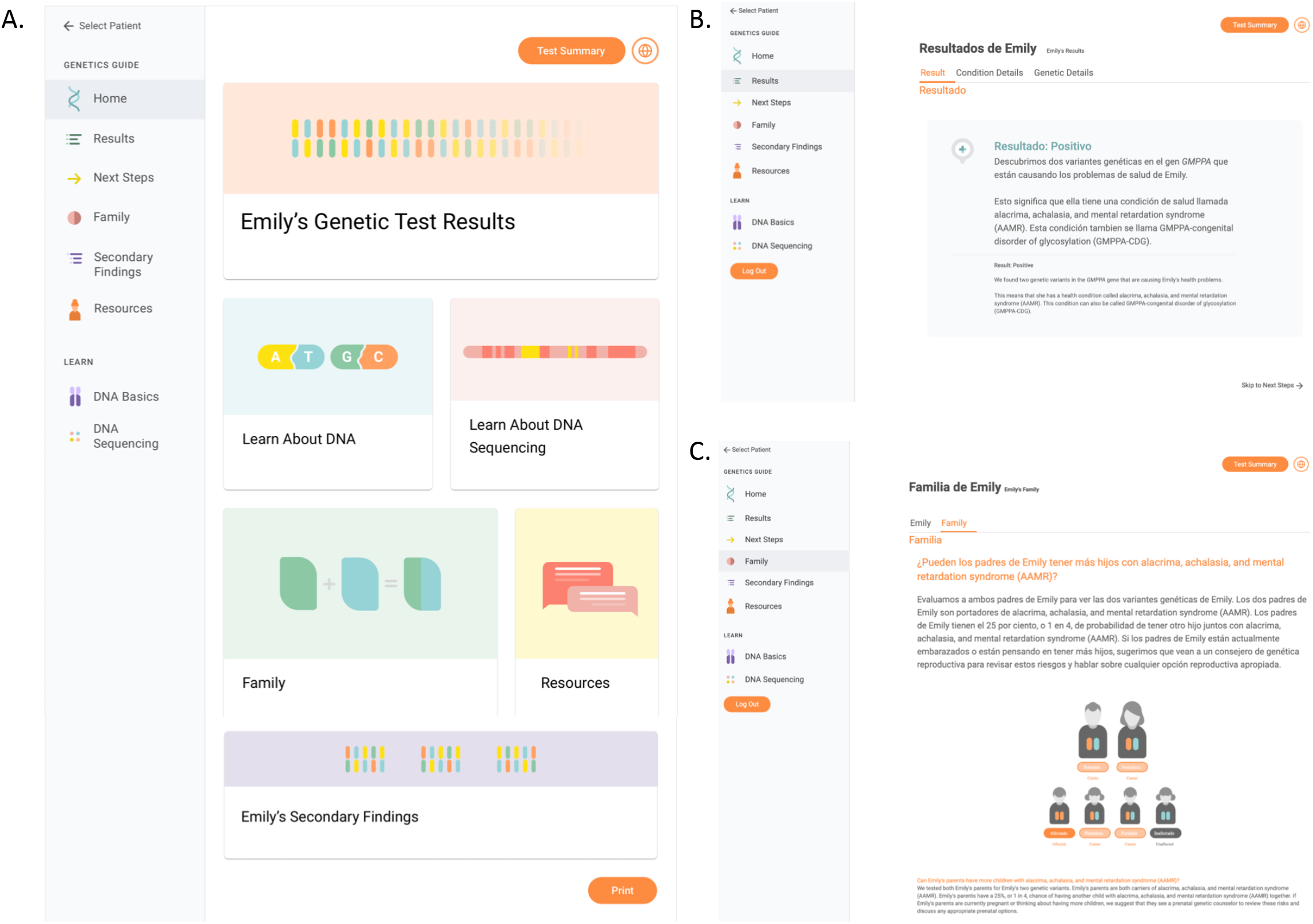
Images of GUÍA web pages showing (A) the GUÍA home page, (B) result summary page displayed in Spanish and English and (C) family page displayed in Spanish and English.

### Evaluating GUÍA During NYCKidSeq Pilot Phase

Eighteen NYCKidSeq pilot phase participants completed feedback interviews, of whom all 18 were female, the mean age was 44 years (range 28-56 years), 2 (11%) were AA, 10 (56%) were H/L, 5 (28%) were EA, and 1 (6%) was MA (**Table 2**). Six (33%) interviews were performed in Spanish. Six (33%) participants’ children received positive genetic results, 3 (17%) received negative results, and 9 (50%) received uncertain results. During the result counseling session, GUÍA was used by both the participant and counselor to guide the delivery of information.

**Table 2.** Socio-demographic characteristics of parent-participants enrolled in the pilot phase of the NYCKidSeq study

| Demographic characteristic (N=18) |  | N (%) |
| --- | --- | --- |
| Sex |  |  |
|  | Female | 18 (100) |
| Age (mean, range) |  | 44 (28 – 56) |
| Self-identified ancestry |  |  |
|  | African American | 2 (11) |
|  | Hispanic/Latinx | 10 (56) |
|  | European American | 5 (28) |
|  | More than one ancestry | 1 (6) |
| Preferred language |  |  |
|  | Spanish | 6 (33) |
| Category of child's primary genomic result |  |  |
|  | Positive | 6 (33) |
|  | Negative | 3 (17) |
|  | Uncertain | 9 (50) |
| Education level |  |  |
|  | Less than High School Graduate | 5 (28) |
|  | High School Graduate | 6 (33) |
|  | Vocational Program | 1 (6) |
|  | Associate College Degree | 3 (17) |
|  | Bachelor's Degree | 2 (11) |
|  | Doctoral Degree | 1 (6) |
| Annual household income |  |  |
| | <\$39,000 | 8 (44) |
| | \$40,000 - \$79,000 | 2 (11) |
| | \$80,000+ | 5 (28) |
|  | Preferred not to answer | 3 (17) |

Prior to meeting with families, GCs built a participant’s case in GUÍA by inputting case details, such as the genetic results, condition-specific details, recommendations for medical management, and patient-participant resources, which are displayed securely on GUÍA via an iPad. Four GCs used GUÍA during the pilot phase. After initial training, GCs report spending approximately 15 minutes inputting data for each case. When asked to describe their experience using GUÍA compared to traditional counseling tools (e.g., flipbooks), GCs highlighted the advantage of having information displayed in both Spanish and English when counseling a Spanish-speaking family using a telephone interpreter. They also expressed that GUÍA provided GCs the ability to personalize patient-participants’ results, saved time in developing patient-participant educational material, and made it easier to communicate results with other providers. Limitations of GUÍA were noted as: restricted to GS tests; children could disrupt the session by their interest in the iPad; patient-participants do not have access to an online version of the tool; and GUÍA does not allow for annotation on pages.

In the feedback interview, participants’ general reactions to GUÍA were overwhelmingly positive (**Table 3**). Specifically, participants felt that GUÍA made receiving information about the result manageable, and they appreciated being able to read along as the counselor discussed the results. Spanish-speaking participants valued reading text in their preferred language. All participants (N=10) who had previous experience with genetic testing agreed that using GUÍA improved result disclosure. Seventeen (94%) participants said the content was clear and all participants stated that the amount of information contained in GUÍA was the right amount. Participants appreciated that they could control the flow and amount of information provided to them.

**Table 3.**
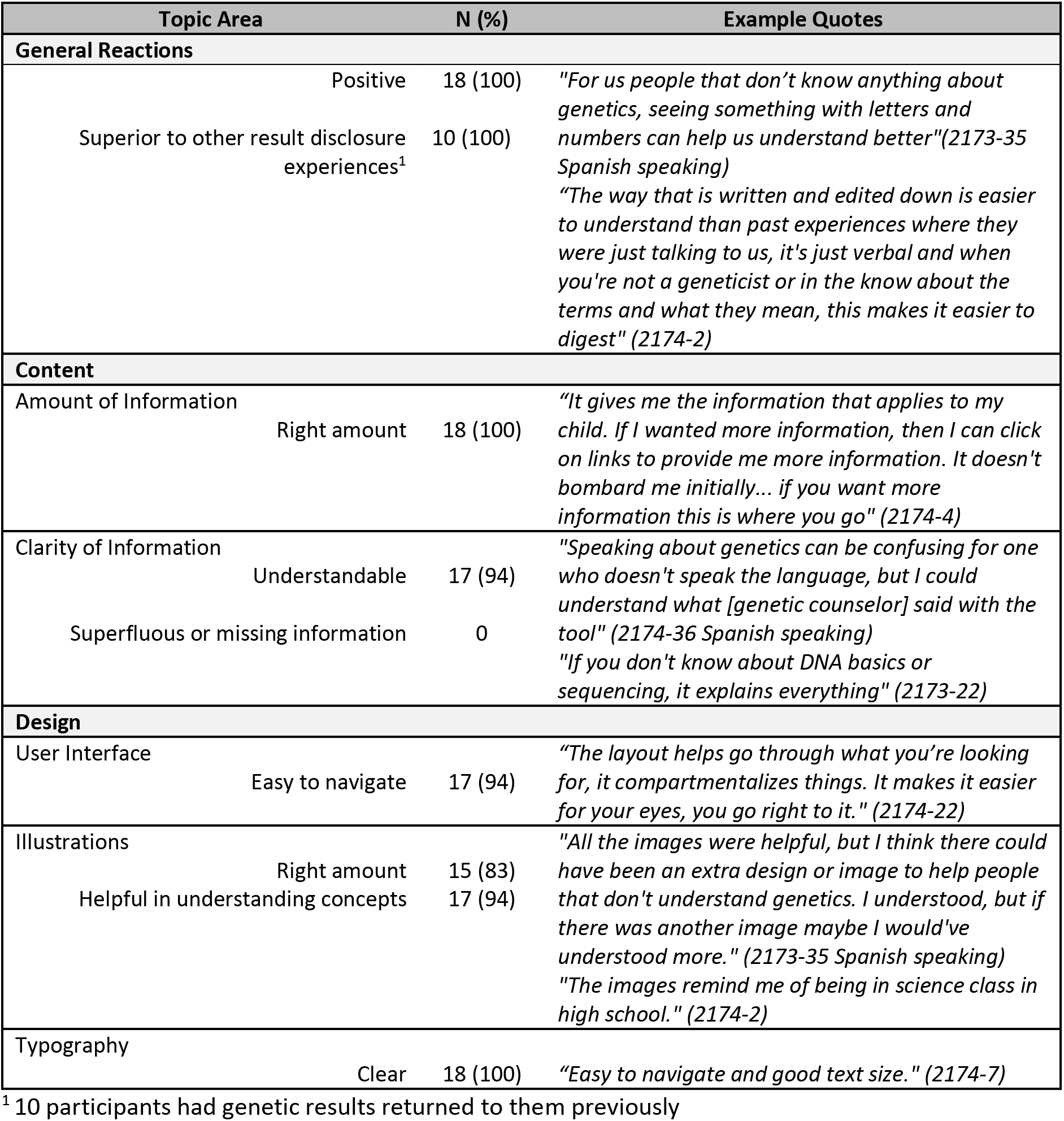
NYCKidSeq parent-participants reactions to GUÍA and feedback on the content and design

Seventeen (94%) participants felt that the user interface was easy to navigate. One participant suggested including a “back” button on every page to better enable the user to navigate between pages of the application. While the majority of participants felt that the number of illustrations included in GUÍA was the right amount and that the illustrations helped them understand complex genetic concepts (15 (83%), 17 (94%), respectively), one participant stated that including additional illustrations would be helpful for participants with limited understanding of genetics.

## DISCUSSION

We describe the design, development process, and evaluation of GUÍA, a novel web-based platform for communicating GS results. Digital health tools like GUÍA have the potential to reduce disparities in access to genomic services by broadening the reach of genetics specialists in diverse and underserved settings and simplifying genomic medicine delivery for non-genetics providers.^23^ GUÍA allows highly technical information to be communicated in an understandable, personalized, and digestible manner. It enables patients to actively engage with their results, control the speed and depth of information delivery, and return to the result information over time. Participants in the NYCKidSeq pilot phase responded positively to the use of GUÍA in their result disclosure session, and all parents with prior genetic testing experience expressed that receiving genomic results with GUÍA was superior. These findings provide preliminary evidence of families’ satisfaction with the integration of this novel application into genetic counseling. The utility of GUÍA to improve parental outcomes will be thoroughly investigated through the NYCKidSeq clinical trial where participants across a variety of clinical settings will be randomized to genetic counseling using GUÍA versus traditional genetic counseling.

The clinical utility of genomic information is inherently linked to understanding of results and adherence to medical recommendations. Traditionally, the communication of clinical results is by genetic specialists, but there is a relative scarcity of medical geneticists and GCs in the United States and worldwide.^24,25^ Proposed methods to scale genomic medicine services include training more genetics professionals and training non-genetics professionals in genomic communication.^26^ Technological solutions can also be leveraged to increase access to genomic medicine services.^6,23^ Research in this space has explored different modalities for scaling genetic counseling services, such as using web-based decisional aids and educational modules for patients to review prior to their appointment.^27^ These efforts have primarily focused on pre-test genetic counseling, likely due to the amount of education typically occurring during the initial appointment, and are not designed to improve patient understanding of their genetic results. GUÍA is well positioned to support the growth of applied GS in that it enhances results disclosure by providing a learning path throughout the genetic counseling process and after. Furthermore, GUÍA can be adapted for a variety of clinical contexts including enhancing telehealth offerings, which is paramount to ensuring broader access and increasing the evidence base for genomic medicine implementation in diverse health care settings.^28^

Currently, the clinical utility of GS tests is limited in historically underserved populations, mainly due to underrepresentation in genomic research and databases.^29-32^ Large-scale research efforts are actively addressing this disparity by expanding genomic databases to better represent diverse populations.^15,33-35^ The fruits of these efforts will increase the frequency with which GS tests provide conclusive results to patients of diverse ancestral backgrounds, and this improvement in clinical utility across populations will further propel the application of GS. To ensure this improvement in accuracy promulgates health equity, equal attention must be paid to accessibility to and communication of genomic health information. Digital communication tools that are culturally aware and multilingual will be needed to best serve diverse communities. Additionally, as the use of GS increases, non-genetics providers are increasingly required and challenged to interpret and discuss GS results with their patients.^36^ We found that non-genetics providers in the NYCKidSeq Program appreciate GUÍA as a provider resource since it deconstructs the clinical report and provides a summary of what was communicated to the family by the GC. Future research could explore the impact of GUÍA on non-genetic provider outcomes and methods for implementing GUÍA to support the results disclosure in non-genetics clinics.

GUÍA was developed by medical professionals within the context of a research program and not in a standard software development environment, which imposes pragmatic limitations on feature development and portability. This could also lead to challenges in sustaining the software beyond the timeline of the project as there are limited resources. GUÍA is HIPAA compliant and patient-participant information is secure, but as GUÍA grows and adapts it will be important to ensure that it remains HIPAA compliant and secure for access outside the health system. For now, GUÍA is only available in English and Spanish, with limited utility to families with a different primary language. In a diverse city like New York, where over 34% of citizens speak a different language in their home, this may curtail the utility of the tool. The pilot evaluation of GUÍA described here was carried out with a small sample and might not be reflective of a broader population.

## Conclusion

To realize the full benefit of genomic medicine, patients and families must understand their genomic results and make informed decisions utilizing this information. Genomic information must be rendered accessible to all individuals regardless of health literacy and English language fluency. Technological solutions will have the greatest positive impact on genomic medicine by reaching as many populations as possible. GUÍA was developed as a bilingual tool to address a large and varied population beginning with the diverse NYCKidSeq participants. Preliminary evidence suggests that GUÍA will be well-received and valued by patients, research participants, families and providers.

## Data Availability

Data is available from the corresponding author upon request.

## Acknowledgements

The authors would like to acknowledge the members of the Genomics Community Board for their valuable input during the development of GUÍA. We would also like to thank Selina Silvas for her contributions to the design of GUÍA.

